# Effect of pre-exposure prophylaxis on risky sexual behaviour of female sex workers in Dakar, Senegal: A randomised controlled trial

**DOI:** 10.1101/2024.08.08.24311660

**Authors:** Wally Toh, Aurelia Lépine, Khady Gueye, Mame Mor Fall, Abdou Khoudia Diop, El hadji Alioune Mbaye, Cheikh Tidiane Ndour, Owen O’donnell

## Abstract

**Background:** HIV prevention through pre-exposure prophylaxis (PrEP) may encourage riskier sexual behaviours that undermine the protection afforded by PrEP and generates negative spillovers on sexually transmitted diseases. Tests for such risk compensatory behaviour in high-risk populations, such as the female sex workers (FSW) in Senegal we studied, are lacking.

**Methods:** We stratified the sample by self-reported sexual risk-taking and prior PrEP experience and randomly assigned them to PrEP referral in October 2021-January 2022 (treatment, n=300) or to deferred PrEP referral after the endline survey in April-May 2022 (control, n=200). We randomised 500 FSWs to start PrEP and included 308 FSWs in the final analysis (T=182, C=126). We compared outcomes in the period preceding PrEP referral of the control group. The primary outcome was self-reported condom use with clients. Secondary outcomes were self-reported sexual risk taking, number of clients, proportion of regular clients, perceived HIV risk of clients, and type of sex act. We estimated intention-to-treat effects of PrEP referral and both average treatment effects on the treated and local average treatment effects of PrEP utilisation. The trial is registered with ISRCTN (ISRCTN16445862).

**Findings:** PrEP referral increased the probability of using PrEP by 34.5 percentage points (pp) [95% CI: 25.4, 43.6; p<0.0001, control group mean: 11.1%]. Estimated effects of PrEP referral and PrEP use on condom use with the last client were 3.3 pp [95% CI: -4.0, 10.6; p=0.376] and 7.9 pp [95% CI: -10.4, 26.3; p=0.397 respectively (control group mean: 84.9%). When looking at condom use with all last three clients, these effects were 11.0 pp [95% CI: 0.8, 21.1; p=0.034] and 25.8 [95% CI: 5.2, 46.4] respectively (control group mean: 67.5%) There were no notable effects on other risky behaviours.

**Interpretation:** This randomised experiment did not give strong grounds for concerns that PrEP encouraged sexual risk-taking by FSWs, at least in the short-term. Robustness of this finding should be tested in larger, longer-term studies and in other contexts. Whether PrEP users are more likely to overreport condom use than non-PrEP users should also be further investigated.

**Funding:** MRC Public Health Intervention Development Scheme from UKRI and D.P. Hoijer Fonds, Erasmus Trustfonds, Erasmus University Rotterdam, The Netherlands.

**Research in context:** *Evidence before this study:* Pre-exposure prophylaxis (PrEP) is becoming an essential part of HIV prevention among high-risk populations in low-and middle-income countries. However, compensatory risky behaviour may partially offset the protection PrEP gives against HIV and may increase the prevalence of other sexually transmitted diseases. Risk compensation may exhibit more strongly among female sex workers (FSWs) as condomless sex is better renumerated. We searched PubMed in January 2024 using the terms (prep) AND (female sex workers) AND (randomised controlled trial) (28 search results) and (PrEP) AND (risk compensation) (140 search results). We did not find any randomised controlled trial testing whether PrEP uptake impacts the prevalence of unprotected sex amongst FSWs. For men who have sex with men, the evidence was mixed, with risk compensation evident in more recent studies.

*Added value of this study:* To our knowledge, this study provides the first evidence from a randomised experiment on whether PrEP causes compensatory risky sexual behaviour by FSWs. We found no strong evidence that either PrEP referral or PrEP use reduces condom use or increases measures of sexual risk-taking. Conversely, we found that PrEP users believe that PrEP is more effective when used with condoms.

*Implications of all the available evidence:* It is generally believed that prescribing PrEP to populations at high risk of HIV infection improves HIV prevention, despite some offsetting risk compensation, but may have a negative impact on the control of other sexually transmitted infections. Our study, which should be replicated with larger samples and in other contexts, suggests that concerns about a negative spillover effect of PrEP on risky sexual behaviour of FSWs may not be well founded. However, this could change if beliefs about dependence of PrEP effectiveness on complementary condom use were to change.

## Introduction

When taken as prescribed, pre-exposure prophylaxis (PrEP) is highly effective in preventing HIV infection [1–2]. However, there is some concern that this protection may encourage risky sexual behaviours — *risk compensation —* that partially offsets the preventive effect [2–7]. For example, those taking PrEP may reduce their use of condoms or increase their number of sexual partners. Such behaviours could increase the prevalence of other sexually transmitted infections (STIs) that PrEP does not protect against.

Most research on behavioural responses to PrEP have focused on men who have sex with men (MSM). Evidence of risk compensation in this population is mixed [2, 8–10]. Risk compensation seems more evident in more recent studies [8–10], which is consistent with an increasing behavioural response as awareness of PrEP’s effectiveness becomes more widespread [8–9].

Female sex workers (FSWs) can charge a higher price for condomless sex and other risky sexual acts. This financial incentive may increase the prevalence of risk compensatory behaviour by FSWs taking PrEP [2, 11]. However, there is a lack of robust evidence on the phenomenon in this population. Observational studies have shown mixed evidence [7, 11, 13–14] that is difficult to interpret because FSWs who take PrEP tend to engage in riskier sexual behaviours and hence, may already be at higher risk of STIs prior to taking PrEP [16]. Qualitative research suggests that the risk of risk compensation may grow as FSWs become more convinced of the effectiveness of PrEP [12]. On the other hand, PrEP provision may also bring hard-to-reach populations, such as FSWs, into closer contact with the health system, increasing access to free condoms and lubricants and improving opportunities for routine STI testing and sexual health counselling [3].

In Senegal, sex work is legal upon registration. Registered FSWs are required to visit a public health centre every month for health checks. Due to the strong stigma attached to sex work, many FSWs choose to remain unregistered. In 2015, HIV/AIDS prevalence among FSWs (6.6 percent) was nine times higher than in the overall population [18]. In 2021/2022, Senegal rolled out PrEP to targeted high-risk populations, including FSWs. Before the rollout, PrEP had been temporarily offered to only a limited cohort of FSWs in a 2015/16 PrEP Demonstration Project, and was later discontinued. Our study aims to use the randomised rollout of PrEP to FSWs in Senegal to obtain the first causal randomised evidence for this high-risk population on risk compensation in response to PrEP.

## Methods

The trial is registered with ISRCTN (registration number: ISRCTN16445862 https://doi.org/10.1186/ISRCTN16445862) and full protocol is available on the UCL repository.

### Study design and participants

The study was a stratified randomised controlled trial. It was approved by the National Ethics Committee for Research and Health Senegal (CNERS) and University College London Research Ethics Committee. Participant consent for survey participation was obtained for each survey.

The sampling frame was a survey of FSWs conducted between June and August 2020 in Dakar, Senegal. The survey was the third wave (after waves in 2015 and 2017) of study following a cohort of FSWs at least 18 years old at entry. In each wave, the sample was replenished with new participants who were recruited via snowball sampling by midwives at public health centres for registered FSWs and by sex workers’ leaders for unregistered FSWs.

We restricted the sample to survey participants who, based on information available in 2020, were potentially eligible for PrEP by excluding those who a) reported not doing sex work in 2020 or in any survey wave were recorded as having a public health system medical record showing they were HIV positive.

### Randomisation

We randomised the survey participants potentially eligible for PrEP to treatment and control groups (3:2). Randomisation was stratified by (a) reported prior experience with PrEP through the 2015/16 PrEP Demonstration Project [24], and (b) sexual risk-taking self-reported in the 2020 survey (Supplementary Material (SM) Text S1).

### Intervention

The intervention was referral of female sex workers to partners in charge of implementing PrEP in Senegal. From October 2021 to January 2022, midwives at public health centres and FSW peer facilitators actively contacted FSWs in the experimental treatment arm and asked if they were interested in receiving it. An appointment to screen for PrEP eligibility was made for those who were interested in PrEP. The Ministry of Health and Social Action (MoH) and a non-governmental organization (NGO) – the Alliance Nationale Contre le SIDA (ANCS) – were the entities responsible for PrEP implementation, which includes eligibility screening, PrEP distribution and follow-up visits. MoH was mainly responsible for registered FSWs, and PrEP screening and distribution took place at public health centres. ANCS mainly targeted unregistered FSWs and used community sites and mobile clinics for PrEP screening and distribution. PrEP eligibility was set by national guidelines and required the potential PrEP candidate to be: a) still doing sex work and b) meeting medical criteria: (i) good liver function measured by creatinine level, (ii) HIV negative, and (iii) not pregnant. Those who eligible were offered PrEP.

During the endline survey in April-May 2022, survey respondents in the experimental control arm who were still in sex work and interested in receiving PrEP were referred for PrEP screening.

### Data collection

As MoH and ANCS were responsible for PrEP implementation, we have limited and incomplete information on the losses at each stage of the recruitment, PrEP uptake and loss-to-follow up in the treatment arm, and no information on the control arm.

Hence, the primary source of data for this analysis comes from the endline survey which was held in April-May 2022. Midwives and FSW peer facilitators attempted to contact all FSWs randomised to either the treatment or control arms. The interviews were carried out by trained enumerators at four public health centres, rented premises nearby, or quarters of trusted FSW leaders. The survey asked about PrEP utilisation, preventive health and sexual behaviours. Self-reported PrEP utilisation during the survey was cross-checked as far as possible with MoH and ANCS.

### Outcomes

The primary outcome was condom use with clients self-reported in the endline survey. We asked each participant to recall their last three clients and whether a condom had been used with each of them. We created two binary outcomes: a) 1 if a condom was used with the last client, and 0 otherwise; b) 1 if a condom was used with all three clients, and 0 otherwise.

The secondary outcomes were other aspects of sexual risk-taking. We asked participants to report on an 11-point Likert scale how much risk they took in their recent sexual behaviours (0 = Limits risks, 10 = Likes to take risks). We also asked participants to report their number of clients in a typical week and the average number of sex acts performed with each of the last three clients. Participants also reported their perceptions of the HIV risk of each of their last three clients (11-point Likert scale: 0 = no risk, 10 = very high risk). We used the maximum of the three observations for each participant as the outcome. In addition, we used the share of clients in a typical week the participant reported to be regulars (as opposed to occasional clients). We also asked each participant to report whether they had oral sex and anal sex with any of their last three clients. Since a low proportion (<1%) reported anal sex with any of their last three clients, this outcome is not reported.

We used a 5-point Likert scale (1: Very unlikely, 5: Very unlikely) to elicit beliefs about the likelihood of contracting HIV (STI) after sexual activity with a person with HIV (STI) under different circumstances: a) with and without use of a condom, b) with no prevention, c) with a condom only, d) with PrEP only, and d) with both PrEP and a condom.

### Pre-experiment power calculation

We calculated the minimum detectable effect (MDE) on condom use given the fixed sample size available from the 2020 survey. Our sampling frame came from a survey of 604 FSWs conducted between June and August 2020 in Dakar, Senegal. We identified 500 respondents who were potentially eligible for PrEP and randomly assigned them into treatment and control groups (3:2). We assumed a drop-out rate of 10% (survey and sex work attrition), a 5% PrEP ineligibility rate, a PrEP adoption rate of 82.4% based on the rate achieved in the 2015/16 PrEP pilot [19], and a baseline condom use rate of 67.9% based on the estimated prevalence in the 2020 survey elicited using list experiments. With 80% power and a two-sided two-sample proportion test (alpha=0.05), we calculated the intention to treat MDE of PrEP referral as 11.9 percentage points and the MDE of PrEP use of 9.3 percentage points. We did not allow for covariates in the power calculations but adjusted for them in the analysis.

### Statistical analyses

We report the proportions of the treatment group and control group who were taking PrEP at the time of the endline survey and who had recently taken PrEP, with binomial exact confidence intervals. We used logistic regression, with robust standard errors, to estimate the effect of assignment to PrEP referral – consisting of screening to establish eligibility and the offer of PrEP if eligible – on the probability of using PrEP.

We estimated intention-to-treat (ITT) effects of PrEP referral on the outcomes. We did not estimate effects of being deemed eligible since we had no information about eligibility in the control arm. Adjusted logistic and ordinary least squares (OLS) regression were used to estimate effects on binary and continuous outcomes, respectively. For each binary outcome, we report the estimated effect on the outcome probability averaged over the sample. Confidence intervals and p-values were obtained from robust standard errors.

To estimate effects of PrEP use on outcomes allowing for non-random variation in use resulting from two-sided noncompliance, we used exogenous variation in use generated by random assignment to PrEP referral. For each binary outcome, we used an adjusted recursive bivariate probit model for use (as a function of referral) and the respective outcome to estimate the average treatment effect of use on the outcome probability. For each continuous outcome, we used two-stage least squares (2SLS), with randomisation to PrEP referral as an instrument utilisation, to estimate the local average treatment effect of use on the respective outcome. In both cases, we used robust standard errors to construct confidence intervals.

Given the limited sample size, we transformed categorical variables into binary indicators of above (1) or below (0) the median: a) self-reported sexual risk (range: 0-10; median: 1), b) perceived client HIV risk (range: 0-10; median: 0), c) beliefs about risk of contracting HIV/STI with protected sex (range: 1-5; median: 1), d) beliefs about risk of contracting HIV/STI with unprotected sex (range:1-5; median: 5). We used logistic regression and recursive bivariate probit to estimate effects of PrEP referral and utilisation, respectively, on these outcomes. In supplementary analysis, we treated these categorical variables as continuous linear measures and used OLS and 2SLS to estimate effects on them. .

Covariates used for adjustment were age in 2022, exposure to Ramadan within 7 days of the 2022 survey interview, and, in 2020, marital status, FSW registration status, self-reported sexual risk taking, prior PrEP experience and the value of the respective outcome in 2020.

To look at how PrEP users perceive the substitutability of PrEP and condoms as HIV preventive methods, we compared whether a higher proportion of PrEP users deemed the risk of contracting HIV to be unlikely when both PrEP and condom are used, as opposed to using just PrEP or just condoms.

All statistical analyses were performed using R 4.1.2, except for recursive bivariate probit models which were estimated in STATA 17.0. This trial is registered with <u>ISRCTN</u> <u>(ISRCTN16445862)</u>. No harm or unintended effects were reported.

## Results

Out of 604 FSWs in the sampling frame (2020 FSW survey participants), 500 were assessed as potentially eligible for PrEP and were randomised to active PrEP referral (treatment, n=300) or delayed (until after the endline survey) PrEP referral (control, n=200) (**Figure 1**). Out of the 500 FSWs randomised, 308 were included in analysis sample, with 95 excluded because they could not be traced or they refused to participate in the survey and 97 excluded because they reported not doing sex work at the time of the survey.

**Figure 1:**
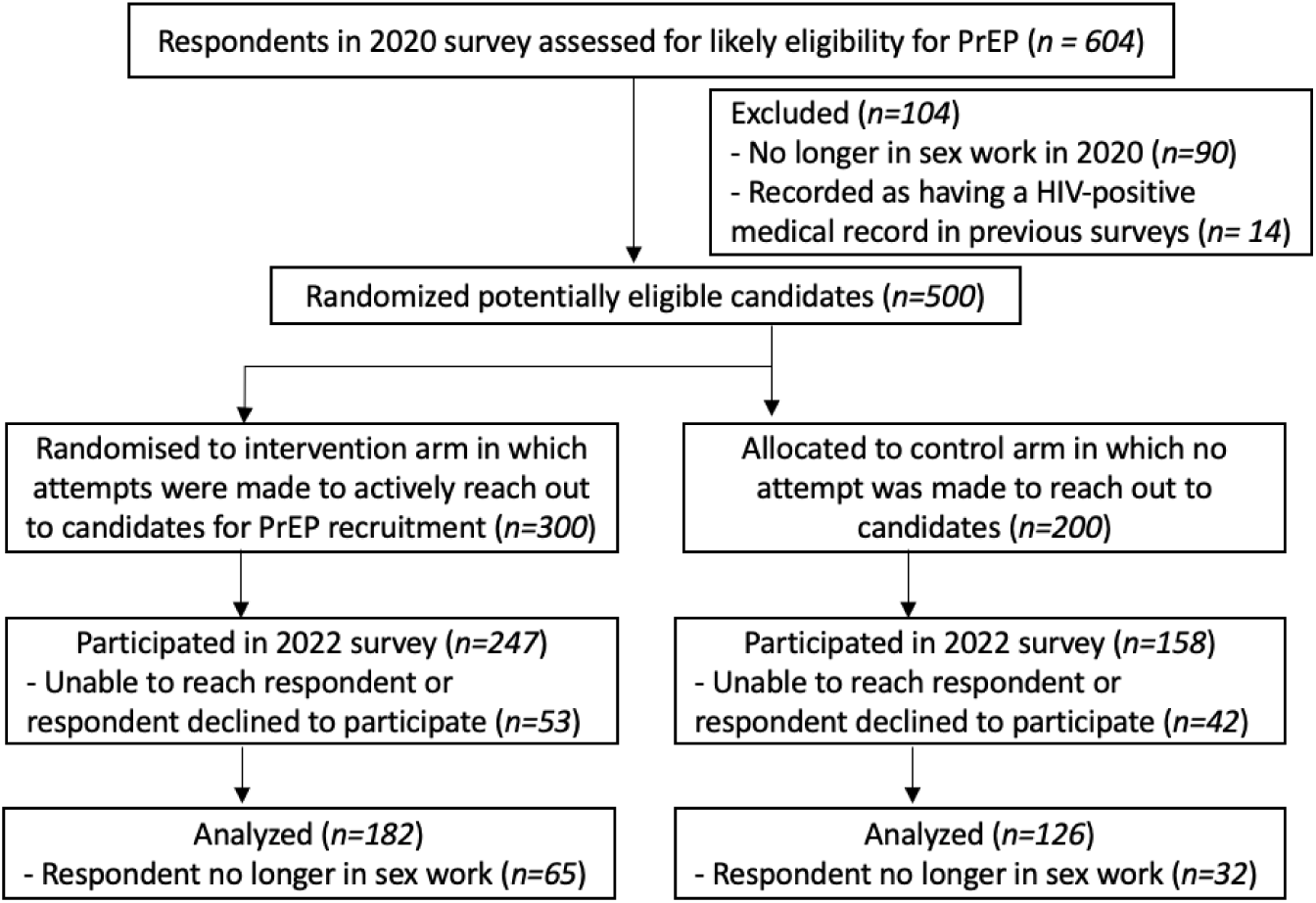
Participant flow.

**Table 1** shows characteristics of the analysis sample at the time of the 2020 survey (baseline), two years before randomisation. Overall, the sample had a mean age of 39 years, over half (54%) had no schooling, almost four fifths (79%) were divorced, separated or widowed, 45% were registered FSWs, a vast majority (97%) reported using a condom with their last client, and only 2% reported using PrEP in 2020. On average, participants had 6.5 clients in a typical week, 72% of their clients were regulars, and they derived 84% of their income from sex work.

**Table 1:** Analysis sample characteristics overall and by treatment status, at baseline in 2020.

|  | Overall<br>(n=308) | Control (C)<br>(n=126) | Treatment (T) (n=182) | p-value<br>(H<sub>0</sub>: C=T) |
| --- | --- | --- | --- | --- |
| **Overall F-test** |  |  |  | 0.275 |
| **Sociodemographics** |  |  |  |  |
| Age (years) | 38.9 (9.4) | 39.2 (9.2) | 38.8 (9.5) | 0.700 |
| Ever went to school |  |  |  | 0.442 |
| Yes | 141 (46%) | 61 (48%) | 80 (44%) |  |
| No | 167 (54%) | 65 (52%) | 102 (56%) |  |
| Marital status |  |  |  | 0.014 |
| Never married | 63 (20%) | 16 (13%) | 47 (26%) |  |
| Married | 3 (1%) | 2 (2%) | 1 (1%) |  |
| Divorced/Separated/<br>Widowed | 242 (79%) | 108 (86%) | 134 (74%) |  |
| Household is indebted |  |  |  | 0.580 |
| Yes | 179 (59%) | 75 (61%) | 104 (58%) |  |
| No | 124 (41%) | 48 (39%) | 76 (42%) |  |
| **Sex work** |  |  |  |  |
| Registered with authorities |  |  |  | 0.569 |
| Yes | 138 (45%) | 54 (43%) | 84 (46%) |  |
| No | 170 (55%) | 72 (57%) | 98 (54%) |  |
| Clients in typical week (No.) | 6.5 (6.0) | 5.6 (4.2) | 7.1 (6.9) | 0.032 |
| Clients in last 7 days (No.) | 2.6 (4.1) | 1.9 (2.1) | 3.0 (5.0) | 0.014 |
| Sex work income in last 7 days ('000<br>CFAF) | 23.1 (52.2) | 19 (24.7) | 25.9 (64.7) | 0.250 |
| Average monthly sex work income ('000<br>CFAF) | 124.5 (105.2) | 125.2<br>(113.1) | 124 (99.8) | 0.928 |
| Average monthly non-sex work income<br>( '000 CFAF) | 24.8 (48.8) | 22.2 (36.1) | 26.6 (55.9) | 0.450 |
| Share of sex work income in total<br>income (Prop.) | 0.84 (0.21) | 0.85 (0.20) | 0.83 (0.21) | 0.473 |
| **Sexual risk-taking** |  |  |  |  |
| Self-reported sexual risk-taking attitude<br>(0=Limits risks, 10=Likes to take risk) |  |  |  | 0.528 |
| Low (<=1) | 140 (45%) | 53 (42%) | 87 (48%) |  |
| High (> 1) | 168 (55%) | 73 (58%) | 95 (52%) |  |
| Condom use with last client |  |  |  |  |
| Yes | 298 (97%) | 120 (96%) | 178 (98%) | 0.359 |
| No | 9 (3%) | 5 (4%) | 4 (2%) |  |
| Share of regular clients in typical week<br>(Prop.) | 0.72 (0.3) | 0.72 (0.30) | 0.71 (0.30) | 0.859 |
| **Randomisation strata** |  |  |  | 0.707 |
| PrEP-ever used + High risk-taking | 34 (11%) | 17 (13%) | 17 (9%) |  |
| PrEP-never used + High risk-taking | 82 (27%) | 32 (25%) | 50 (27%) |  |
| PrEP-ever used + Low risk-taking | 67 (22%) | 26 (21%) | 41 (23%) |  |
| PrEP-never used + Low risk-taking | 125 (41%) | 51 (40%) | 74 (41%) |  |

Table shows mean (SD) for continuous/integer variables and n (%) for binary/categorical variables. p-values are from two-sided t-tests of equality of means and proportions for continuous and binary variables, respectively, and from F-tests for variables with more than 2 categories. See SM Text S1 for definitions of randomization strata.

With the exception of differences in marital status and client numbers, the treatment and control groups were balanced on baseline characteristics. A joint F-test did not reject the null hypotheses of no differences across the multiple characteristics (p-value=0.275). There was balance in the pre-attrition sample (SM Table S1), indicating that randomisation generated observationally equivalent treatment and control groups (p-value=0.812). Selection into the analysis sample (n=308) from the pre-attrition sample (n=500) did not different by treatment assignment and most baseline characteristics, with the exceptions of household indebtedness and one of the randomisation strata (SM Table S2).

Sociodemographic and sex work characteristics of the overall sample reported at endline (2022) were broadly similar to those reported at baseline (2020) (SM Table S3). There was a 10 percentage point (pp) reduction in reported condom use with the last client, but a higher proportion who reported limiting sexual risk-taking.

In the control group stated 11.1% (95% CI: 6.2-7.9) reported using PrEP at the time of the endline survey and 15.9% (95% CI: 10.0, 23.4) reported that they had recently used PrEP. The respective percentages in the treatment group were 45.6% (95% CI: 38.2, 53.1) and 54.9% (95% CI: 37.7, 52.6). Assignment to active PrEP referral increased the probability of currently using PrEP by 34.5 pp (95% CI: 25.4, 43.6; p<0.0001) and the probability of having used PrEP in 2021/22 by 39.1 pp (95% CI: 29.4, 48.7; p<0.0001).

Table 2 shows estimates of ITT effects of PrEP referral, effects of PrEP use on condom use and effects of having used PrEP in 2021/2022. The latter two differ in that some respondents may have started using PrEP but discontinued their usage by the endline survey. These respondents would be recorded as not using PrEP currently but have used PrEP in 2021/2022. The estimated effect of PrEP referral on the probability of using a condom with the last client is 3.3 pp (95% CI: -4.0, 10.6; p=0.376). The point estimate corresponds to a 3.8% increase over the control group mean (84.9%). PrEP referral was estimated to increase the probability of using a condom with all last three clients by 11.0 pp (95% CI: 0.8, 21.2; p=0.034) – a 16.3% increase relative to the control group mean (67.5%). The estimated effect of PrEP use on condom use with the last client is 7.9pp (95% CI: -10.4, 26.3; p=0.397). PrEP use was estimated to increase the probability of using a condom with all of the last three clients by 25.8 pp (95% CI: 5.2, 46.4; p=0.014), which corresponds to a 38.2% increase. Effects of having used PrEP in 2021/2022 were very similar to that of PrEP use.

**Table 2:** Estimated risk difference (RD) in condom use, PrEP referral, PrEP use and ever used PrEP in 2021/22.

|  | Control group | PrEP referral (n = 308) |  |  | PrEP use (n = 308) |  |  | Ever used PrEP in 2021/22 (n = 308) |  |  |
| --- | --- | --- | --- | --- | --- | --- | --- | --- | --- | --- |
|  | mean | RD | 95% CI | p | RD | 95% CI | p | RD | 95% CI | p |
| Condom used with last client (0, 1) | 0.849 | 0.033 | [-0.04, 0.106] | 0.376 | 0.079 | [-0.104, 0.263] | 0.397 | 0.074 | [-0.093, 0.240] | 0.386 |
| Condom used with all last 3 clients (0, 1) | 0.675 | 0.110 | [0.008, 0.212] | 0.034 | 0.258 | [0.052, 0.464] | 0.014 | 0.243 | [0.056, 0.429] | 0.011 |

*Notes:* Risk differences caused by PrEP referral estimated by adjusted logistic regression. Risk differences caused by PrEP use were estimated by recursive bivariate probit with PrEP use instrumented with random assignment to PrEP referral. Adjustment for age, an indicator of Ramadan within the last 7 days, marital status in 2020, FSW registration, self-reported sexual risk taking, prior PrEP experience and the 2020 value of the outcome.

Table 3 shows estimates of effects of PrEP referral and PrEP use on secondary outcomes. With one exception, the 95% confidence intervals all include zero, and the magnitude of the estimates relatively small compared to the control group means. The exception is the estimated effect of PrEP use on the probability of perceiving at least one of the last three clients as having a high risk of being HIV positive. However, this finding is not robust to using an alternative outcome specification (see SM Table S4), while the nonsignificant findings for the other secondary outcomes are robust to alternative outcome specifications.

**Table 3:**
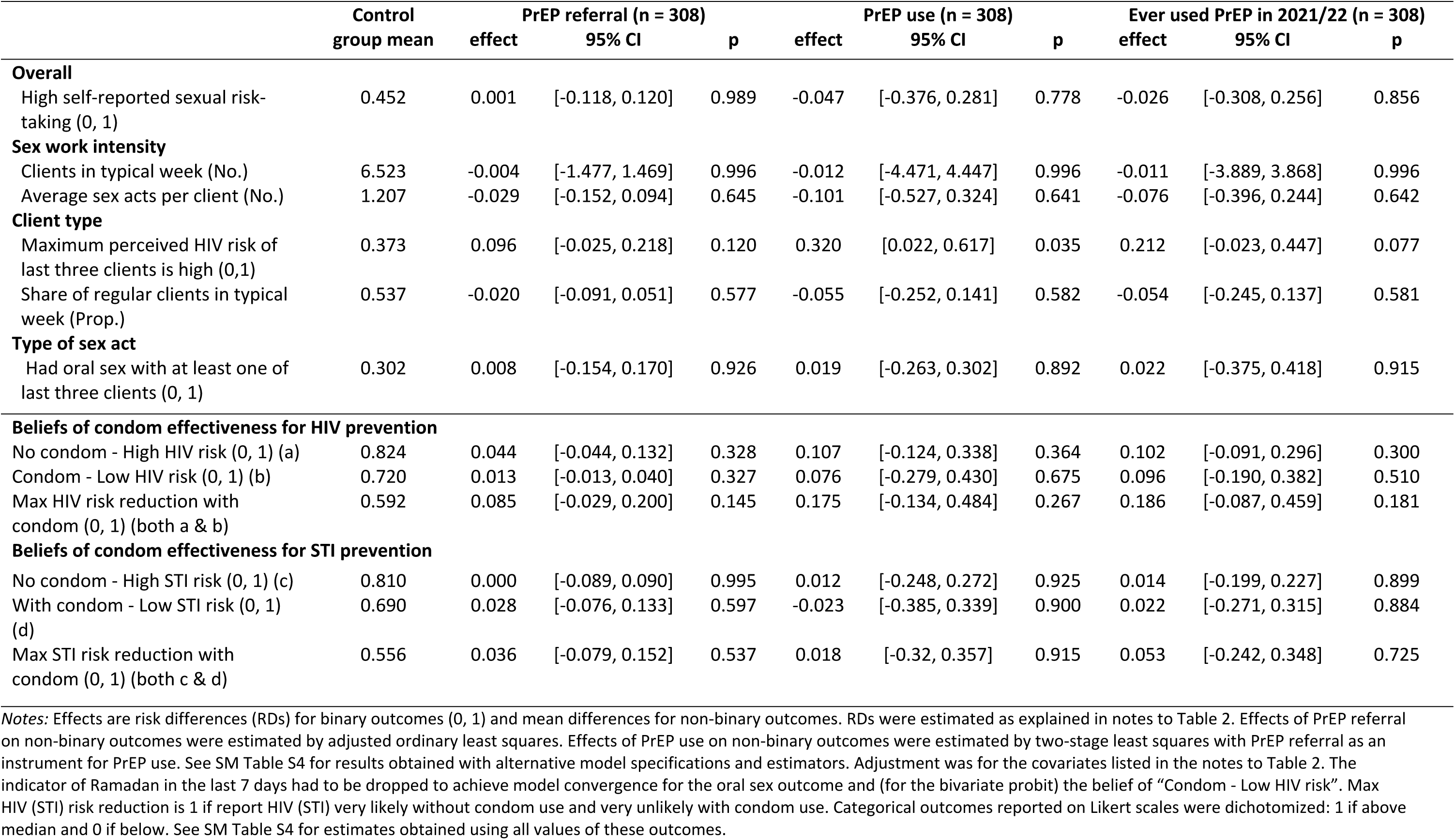
Estimated effects of active PrEP referral and PrEP use on secondary outcomes.

Table 4 shows estimates of effects of PrEP referral and PrEP use on beliefs about the effectiveness of condoms in preventing HIV and STIs. The signs of the point estimates are consistent with PrEP referral and use increasing beliefs that condoms are effective in preventing HIV, although the 95% confidence intervals all include zero. In the treatment group, a higher percentage of participants reported that the likelihood of contracting HIV was (a) very likely if no condom was used (4.4 pp, 95% CI: -4.4, 13.2), (b) very unlikely if a condom was used (1.3 pp, 95% CI: -1.3, 4.0). and (c) both very likely if no condom was used and very unlikely if a condom was used (8.5pp, 95% CI: -2.9, 20.0). The directions of these differences are robust to using the full range of perceived likelihoods of contracting HIV, not just the extremes, and the p-values are smaller in that case (SM Table S5). Estimates of effects of PrEP referral on beliefs about the effectiveness of condoms in preventing other STIs do not show the same consistency, have much larger p-values and much smaller effects.

**Table 4:** PrEP user beliefs about likelihood of HIV infection after sex with HIV+ person (n=97)

|  | Use neither<br>PrEP nor<br>condom | Use only PrEP | Use only condom | Use both PrEP<br>and condom |
| --- | --- | --- | --- | --- |
| Very unlikely | 6 (6%) | 37 (38%) | 39 (40%) | 88 (91%) |
| Unlikely | 0 (0%) | 23 (24%) | 31 (32%) | 5 (5%) |
| Equally likely as unlikely | 0 (0%) | 17 (18%) | 18 (19%) | 1 (1%) |
| Likely | 14 (14%) | 15 (15%) | 7 (7%) | 0 (0%) |
| Very likely | 77 (79%) | 5 (5%) | 2 (2%) | 3 (3%) |

*Notes: Table shows sample frequencies (%). Sample restricted to participants who reported currently using PrEP at time of endline survey in both the treatment and control groups.*

Among PrEP users, when neither PrEP nor condoms were used for protection, 6% of respondents responded that it was very unlikely that they would be infected by HIV after engaging in a sex act with a HIV-positive sexual partner (Table 4). This proportion increased to 38% and 40% when either PrEP or condoms were used respectively. When both PrEP and condoms were used, 91% of PrEP users stated that it was very unlikely they would be infected by HIV. A higher proportion of PrEP users believed condoms to be a more effective HIV prevention method than PrEP: 9% of PrEP users answered that they are likely or very likely to being infected by HIV when using PrEP only versus 20% for condom only.

## Discussion

Our study did not find any experimental evidence of substitution between PrEP and condom use. There was no statistically significant decrease in condom use with the last client with the PrEP referral intervention among PrEP users and FSWs recently exposed to PrEP. When taking all three clients together, we observed an increase in condom use in the treatment group. In addition, we did not observe a notable increase in other risky sexual behaviours.

While we have focused on PrEP usage and recent PrEP uptake in our exposition, it is important to note that PrEP is never offered in isolation, and it may be equally important to consider that the way PrEP is being implemented within the health system may influence outcomes. First, the PrEP programme could potentially allow the health system to gain an additional touchpoint to improve the sexual health and influence the sexual behaviours of this hard-to-reach sex worker population. In the current programme implementation in Senegal, it was emphasized to PrEP users that condoms needed to be used with PrEP as PrEP does not protect against other STIs. In addition, free condoms were also distributed at the recruitment sites part of the registration policy for FSWs in place in Senegal since 1969. Anecdotally, a notable reduction in condom supplies of the MoH was observed after the introduction of PrEP, consistent with the increase in condom use found in the study. While not implemented yet in Senegal, coupling STI screenings among PrEP users may potentially increase the benefits of PrEP, while mitigating the consequences of any risk compensation. Further research on the optimal frequency of STI screenings for this population would be needed [13]. Second, PrEP programme implementation could affect how target users perceived PrEP and influenced the take-up and adherence to PrEP. In our case, we observed that registered FSWs exhibited a higher willingness to initiate PrEP than non-registered FSWs (SM Table S5).

Our estimates on beliefs showed that beliefs on effectiveness of condom use at preventing HIV had not decreased after PrEP uptake. While not statistically significant at conventional levels, the consistency of our estimates robust to changes in model specification suggest that beliefs in condom effectiveness in HIV prevention may even have increased. This is further supported by evidence that a significant proportion of PrEP users believed that condom use provides substantial additional protection to PrEP, and vice versa. Together with an increase in condom use, these pieces of evidence are consistent with the hypothesis that users currently see PrEP as complementary HIV prevention strategy and not a substitute to condom use.

Nonetheless, with respect to the goal of HIV prevention, it is unclear the extent to which condoms are indeed complementary to PrEP from the perspective of an individual user. A substantial decrease in condom use in the population may increase STIs, and having a STI increases the chance of an individual getting HIV. Hence, condom use may still have impact on preventing HIV transmission especially in the general population. However, for the individual user, PrEP is also highly effective if taken as prescribed [1], suggesting that condoms may in fact offer smaller marginal benefit in HIV prevention with consistent PrEP usage than is currently perceived by users. Consequently, there may be a future risk of a downward shift in perception of the marginal benefit of condoms for HIV prevention under consistent PrEP usage by PrEP users. As trust in PrEP as an effective HIV prevention method increases in the longer term, condom usage may potentially decline as a response to these adjusted beliefs.

There are several limitations to our study. First, our intervention involved the investment of substantial effort to trace and recruit FSWs in the treatment arm. It is likely that some of those who started PrEP in the treatment arm might not have been reached and made aware of PrEP. In addition, PrEP acceptability may be a larger issue under business-as-usual recruitment [20], as less time and effort may be invested in counselling and convincing potential users of the benefits of PrEP. Therefore, the proportion of PrEP users in our treatment arm is likely to be higher than expected, and our experimental results may not extend fully to business-as-usual circumstances.

Second, our study is unable to study longer-term outcomes due to various feasibility constraints. Even though the overall HIV prevalence in Senegal is relatively low (0.3%), that of FSWs is still high (4.8%) [21]. Hence, limiting recruitment efforts cannot be sustained long term due to ethical reasons. Furthermore, compliance in the treatment arm is likely to decrease considerably over time, while contamination in the control arm is likely to increase over time, reducing the differences in uptake between the treatment and control arms. In addition, already high attrition from sex work and the survey will worsen with time. Short-term outcomes could differ from longer-term ones, especially since PrEP users in our study seem to hold the perception that condom use still provides substantial additional HIV protection on top of PrEP, even when PrEP should offer 99% protection against HIV when used consistently. Furthermore, the increasing awareness of clients about the introduction of PrEP in Senegal coupled with the low bargaining power of FSWs will likely reduce condom use [11]. In addition, there could also be community-level risk compensation of non-PrEP users when awareness and the prevalence of PrEP usage increases among FSWs [22].

Third, data used in our study is primarily self-reported. Self-reported measures may differ from actual behaviour due to intentional (e.g. due to social desirability bias) and unintentional misreporting (e.g. recall bias). If the degree of misreporting differs between the treated and the control group because of the intervention, the estimates may be biased. In our survey, we employed two other indirect elicitation methods-a double list experiment [23–24], and the colorbox method [25]. The double list experiment used two lists of statements to elicit the prevalence of a sensitive statement (condom use). However, it was unsuitable for this analysis due to its high standard errors and prevalence estimates differed substantially between the two lists. Nonetheless, it provides indications that there may be a divergence between indirect elicitation and direct elicitation in the overall analysis sample as condom use prevalence with the last client was 84.9% [95% CI: 78.6, 91.2] with direct elicitation, but 69.4% [95% CI: 59.5%, 80.1%] with the double list experiment. The latter method which indirectly elicited identifiable individual responses did not work as expected at reducing misreporting as it garnered similar condom use prevalences as direct questioning. Therefore, we used directly elicited condom use in our study. Hence, future studies should further investigate whether PrEP programmes also influence the incentives to misreport condom use.

Fourth, the results from this study must be confirmed in larger studies or use as part of meta-analyses with other studies. Actual PrEP take-up rate was much lower than that the 82.4% seen in Senegal’s 2015 PrEP demonstration study among eligible participants [19]. In addition, the PrEP rollout was originally scheduled to start soon after the 2020 survey to reduce attrition. However, it encountered multiple delays and was eventually rolled out at different times by the two local partners. These reasons contributed to a low two-sided compliance rate and high survey and sex work attrition. Consequently, the study was eventually underpowered.

Fifth, sex work is legal in Senegal and the system already has existing public health care clinics providing care to registered FSWs. In addition, HIV prevalence in Senegal is relatively low. Therefore, the findings of this study may not necessarily extend to countries with different circumstances.

In conclusion, our study suggested no evidence of risk compensation in the short term. In addition, condom use was observed to increase, suggesting that the PrEP programme might have reached FSWs who might have otherwise had lower access to free condom use provision or interface with the health system. Findings suggest that beliefs of condom effectiveness may potentially have increased, and condoms are still viewed as complementary to PrEP usage. Larger studies are required to confirm the findings from this study. Studies on longer term outcomes are also necessary as risk compensation may potentially increase with increase in PrEP usage experience among PrEP users.

## Data Availability

All files are available from the UCL repository database.

## References

[1] PrEP effectiveness [Internet]. Centers for Disease Control and Prevention (CDC). 2022 [cited 2023Jan26]. Available from: https://www.cdc.gov/hiv/basics/prep/prep-effectiveness.html

[2] Murchu EO, Marshall L, Teljeur C, Harrington P, Hayes C, Moran P, Ryan M. Oral pre-exposure prophylaxis (PrEP) to prevent HIV: a systematic review and meta-analysis of clinical effectiveness, safety, adherence and risk compensation in all populations. BMJ open. 2022 May 1;12(5):e048478.

[3] Rojas Castro D, Delabre RM, Molina JM. Give PrEP a chance: moving on from the “risk compensation” concept. Journal of the International AIDS society. 2019 Aug;22:e25351.

[4] Peltzman S. The effects of automobile safety regulation. Journal of political Economy. 1975 Aug 1;83(4):677–725.

[5] Wilson NL, Xiong W, Mattson CL. Is sex like driving? HIV prevention and risk compensation. Journal of Development Economics. 2014 Jan 1;106:78–91.

[6] Carlo Hojilla J, Koester KA, Cohen SE, Buchbinder S, Ladzekpo D, Matheson T, Liu AY. Sexual behavior, risk compensation, and HIV prevention strategies among participants in the San Francisco PrEP demonstration project: a qualitative analysis of counseling notes. AIDS and Behavior. 2016 Jul;20:1461–9.

[7] Giguère, K., Béhanzin, L., Guédou, F. A., Talbot, D., Leblond, F. A., Goma-Matsétsé, E., … & Alary, M. (2019). PrEP use among female sex workers: no evidence for risk compensation. Journal of acquired immune deficiency syndromes (1999), 82(3), 257.

[8] Powell VE, Gibas KM, DuBow J, Krakower DS. Update on HIV preexposure prophylaxis: Effectiveness, drug resistance, and risk compensation. Current infectious disease reports. 2019 Aug;21(8):1–8.

[9] Traeger MW, Schroeder SE, Wright EJ, Hellard ME, Cornelisse VJ, Doyle JS, Stoové MA. Effects of pre-exposure prophylaxis for the prevention of human immunodeficiency virus infection on sexual risk behavior in men who have sex with men: a systematic review and meta-analysis. Clinical Infectious Diseases. 2018 Aug 16;67(5):676–86.

[10] Yan X, Jia Z, Zhang B. Evaluating the risk compensation of HIV/AIDS prevention measures. The Lancet Infectious Diseases. 2022 Apr 1;22(4):447–8.

[11] Quaife M, Vickerman P, Manian S, Eakle R, Cabrera-Escobar MA, Delany-Moretlwe S, Terris-Prestholt F. The effect of HIV prevention products on incentives to supply condomless commercial sex among female sex workers in South Africa. Health economics. 2018 Oct;27(10):1550–66.

[12] Kayesu I, Mayanja Y, Nakirijja C, Machira YW, Price M, Seeley J, Siu G. Uptake of and adherence to oral pre-exposure prophylaxis among adolescent girls and young women at high risk of HIV-infection in Kampala, Uganda: A qualitative study of experiences, facilitators and barriers. BMC women’s health. 2022 Dec;22(1):1–4.

[13] Bowring AL, Ampt FH, Schwartz S, Stoové MA, Luchters S, Baral S, Hellard M. HIV pre-exposure prophylaxis for female sex workers: ensuring women’s family planning needs are not left behind. Journal of the International AIDS Society. 2020 Feb;23(2):e25442.

[14] Mboup A, Béhanzin L, Guédou FA, Geraldo N, Goma-Matsétsé E, Giguère K, Aza-Gnandji M, Kessou L, Diallo M, Kêkê RK, Bachabi M. Early antiretroviral therapy and daily pre-exposure prophylaxis for HIV prevention among female sex workers in Cotonou, Benin: a prospective observational demonstration study. Journal of the International AIDS Society. 2018 Nov;21(11):e25208.

[15] Guest G, Shattuck D, Johnson L, Akumatey B, Clarke EE, MacQUEEN KM. Changes in sexual risk behavior among participants in a PrEP HIV prevention trial. Sexually transmitted diseases. 2008 Dec 1>:1002–8.

[16] Grant RM, Anderson PL, McMahan V, Liu A, Amico KR, Mehrotra M, Hosek S, Mosquera C, Casapia M, Montoya O, Buchbinder S. An observational study of preexposure prophylaxis uptake, sexual practices, and HIV incidence among men and transgender women who have sex with men. The Lancet. Infectious diseases. 2014 Sep;14(9):820.

[17] Senegal [Internet]. Unaids.org. [cited 2023 Dec 2]. Available from: https://www.unaids.org/en/regionscountries/countries/senegal

[18] APAPS & IRESSEF (2016). Enquête nationale de surveillance combinée desinfections sexuallement transmissibles et du VIH/SIDA, Groupe cible: Travailleuses du sexe. Dakar, Senegal.

[19] Sarr M, Gueye D, Mboup A, Diouf O, Bao MD, Ndiaye AJ, Ndiaye BP, Hawes SE, Tousset E, Diallo A, Jones F. Uptake, retention, and outcomes in a demonstration project of pre-exposure prophylaxis among female sex workers in public health centers in Senegal. International journal of STD & AIDS. 2020 Oct;31(11):1063–72.

[20] Witte SS, Filippone P, Ssewamala FM, Nabunya P, Bahar OS, Mayo-Wilson LJ, Namuwonge F, Damulira C, Tozan Y, Kiyingi J, Nabayinda J. PrEP acceptability and initiation among women engaged in sex work in Uganda: Implications for HIV prevention. EClinicalMedicine. 2022 Feb 1;44:101278.

[21] Senegal [Internet]. UNAIDS. 2022 [cited 2022Dec24]. Available from: https://www.unaids.org/en/regionscountries/countries/senegal

[22] Holt M, Murphy DA. Individual versus community-level risk compensation following preexposure prophylaxis of HIV. American journal of public health. 2017 Oct;107(10):1568–71.

[23] Blair G, Imai K. Statistical analysis of list experiments. Political Analysis. 2012;20(1):47–77.

[24] Lépine A, Treibich C, Ndour CT, Gueye K, Vickerman P. HIV infection risk and condom use among sex workers in Senegal: evidence from the list experiment method. Health Policy and Planning. 2020 May;35(4):408–15.

[25] Lépine A, Toh WQ, Treibich C. Colorbox: a novel method for eliciting sensitive behaviours in face-to-face interviewer-led surveys. Unpublished.

